# Healthcare workers’ perceptions and medically approved COVID-19 infection risk: understanding the mental health dimension of the pandemic. A German hospital case study

**DOI:** 10.1101/2022.03.28.22273029

**Authors:** Ellen Kuhlmann, Georg M. N. Behrens, Anne Cossmann, Stefanie Homann, Christine Happle, Alexandra Dopfer-Jablonka

## Abstract

**Introduction:** This study analyses how healthcare workers (HCWs) perceived risks, protection and preventive measures during the COVID-19 pandemic in relation to medically approved risks and organisational measures. The aim is to explore ‘blind spots’ of pandemic protection and make mental health needs of HCWs visible.

**Methods:** We have chosen an ‘optimal-case’ scenario of a high-income country with a well-resourced hospital sector and low HCW infection rate at the organisational level to explore governance gaps in HCW protection. A German multi-method hospital study at Hannover Medical School served as empirical case; document analysis, expert information and survey data (n=1163) were collected as part of a clinical study into SARS-CoV-2 serology testing during the second wave of the pandemic (November 2020-February 2021). Selected survey items included perceptions of risks, protection and preventive measures. Descriptive statistical analysis and regression were undertaken for gender, profession and COVID-19 patient care.

**Results:** The results reveal a low risk of 1% medically approved infections among participants, but a much higher mean personal risk estimate of 15%. The majority (68.4%) expressed ‘some’ to ‘very strong’ fear of acquiring infection at the workplace. Individual protective behaviour and compliance with protective workplace measures were estimated as very high. Yet only about half of the respondents felt strongly protected by the employer; 12% even perceived ‘no’ or ‘little’ protection. Gender and contact with COVID-19 patients had no significant effect on the estimations of infection risks and protective workplace behaviour, but nursing was correlated with higher levels of personal risk estimations and fear of infection.

**Conclusions:** A strong mismatch between low medically approved risk and personal risk perceptions of HCWs brings stressors and threats into view, that may be preventable through better information and risk communication and through investment in mental health and inclusion in pandemic preparedness plans.

## Introduction

The COVID-19 pandemic put a spotlight on the importance of healthcare workers (HCWs) and their contribution to health system resilience. Investment in the health workforce and prioritisation of HCW needs in health policy and pandemic recovery plans is therefore called for by WHO (1,2), the European Commission (EC) (3,4), public health organisations (5) and researchers (6,7), including greater attention to the mental health requirements of the health workforce (8,9,10,11,12,13). Data gathered during the pandemic in different regions of the world revealed a high risk of COVID-19 infection and death among HCWs, as well as an increase in stress and burn-out syndromes in many health professional groups (14,15,16,17,18,19,20,21,22,23,24,25,26,27,28,29). Individual stories of HCWs furthermore illustrate the severity of mental health risks and extremely high workload (30,31).

Lack of attention to the health and wellbeing of HCWs (32) directly impacts in health workforce recruitment and retention (33,34) and weakens health workforce resilience. Every new wave of COVID-19 increased the pressures on HCWs and worsened the health labour market situation and the delivery of patient care. However, the intersecting dynamics and their negative effects on pandemic preparedness and health system resilience are not well understood. As Bourgeault et al. reported in the first year of the COVID-19 pandemic, ‘[P]andemic response plans in country after country, often fail to explicitly address health workforce requirements and considerations’ (35). In year 2, investment in the health workforce ranked still low on the agenda of national ‘coronavirus politics’ (36), but some change can now be observed, as developments in Europe show.

A recent Companion Report, a joint project of the European Commission, the OECD on the European Observatory on Health Systems and Policies, mentioned the future health workforce as one out of three health policy priorities (3,37). The recommendations highlighted that ‘more detailed information on the impact of COVID-19 on health workers’ wellbeing’ is ‘crucial to designing better support measures’ (3:p.31). Similarly, the European Union Expert Panel (38), as well as scholarly debate into building back better after the pandemic mentioned the important role of the health workforce for health systems (4,30,39,40,41,42,43).

EU expert recommendations and international scientific evidence do not easily translate into policy changes in the member states. Health workforce development is still poorly developed and focused on planning and surge capacities. ‘The ‘human face’ (44), the individual person behind every HCW and their wellbeing and needs, is rarely considered. A comparative assessment of HCW protection and preparedness in selected European Union (EU) countries revealed, that ‘action has been taken to improve physical protection, digitalisation and prioritisation of healthcare worker vaccination, whereas social and mental health support programmes were weak or missing’ (12). The findings raise the question, whether health workforce policy and the COVID-19 pandemic protection measures effectively support the needs and requirements of HCWs.

This study seeks to analyse HCWs’ estimations of the personal likelihood of having acquired COVID-19 (infection risk), their fear of infection and their perceptions of protection and preventive measures during the COVID-19 pandemic in relation to medically approved risks and institutional conditions. We have chosen an ‘optimal-case’ scenario of a high-income country with a very well-resourced hospital sector (3,45,46,47) and a low HCW infection rate at the regional (state of Lower Saxony) and the organisational level (48,49) compared to other European countries (16). This research design provides opportunity for exploring the relationship between medically approved risks and physical protection – the ‘objective’ realities and organisational conditions – and the individual perceptions of HCWs, the ‘subjective’ realities and mental health conditions. We aim to reveal ‘blind spots’ of contemporary health workforce governance and COVID-19 pandemic protection, arguing the need for new participatory approaches that integrate the perceptions of HCWs and pay greater attention to mental health requirements.

## Methods

We use Hannover Medical School (50) as an explorative case study, a large university hospital (academic medical centre) in Lower Saxony, a state in the Western part of Germany. Theoretically, our analysis is informed by multi-level health workforce governance (51,52). More specifically, our approach places the perceptions of HCWs in the context of institutional conditions, taking system, sector and organisational levels into account (12). Empirically, we draw on material from the COVID-19 Contact (CoCo) Study, a multi-method study carried out at Hannover Medical School. The study comprised SARS-CoV-2 serology testing of HCWs with patient contact in low-prevalence settings and an additional questionnaire-based online survey; inclusion criteria were all HCWs working in patient care or in units with possible COVID-19 contact, e.g. emergency services (Box 1).

For the purpose of our analysis, we used survey data gathered during November 2020 to February 2021, covering the second wave of the COVID-19 pandemic in Germany. Hospital HCWs (n=1163 respondents) were defined as all persons employed by Hannover Medical who met the inclusion criteria, regardless of their profession/occupation (53) (see Box 1). Selected items of the questionnaire included perceptions of COVID-19 risk, fear, protection and preventive measures (CoCo 2.0, Fragebogen 2; available in German; selected items translated):

- How do HCWs estimate their personal likelihood of having acquired a COVID-19 infection and their fear of infection (items 26, 33, 34)?
- How do HCWs estimate their compliance with infection control and prevention in their private sphere (items 37)?
- How do HCWs perceive the prevention measures at the workplace (items 41, 42)?

Descriptive statistical analysis was undertaken and additional single regression for the items ‘gender’ (female/male), ‘profession/occupation’ (grouped into three categories: ‘physicians’, ‘nurses’, ‘others’) and ‘COVID-19 patient care’ (grouped into two categories: ‘not/not fully protected’, ‘fully protected’) (SPSS®, version 26). In addition, expert information and document analysis (websites, organisational statistics, statements, etc.) were considered to explore the institutional conditions and the organisation’s pandemic management and HCW protection measures.

**Box 1. The COVID-19 Contact (CoCo) Study**

The CoCo Study is an ongoing, prospective, longitudinal, observational study in healthcare professionals/workers and individuals with potential contact to SARS-CoV-2, aiming to improve data and knowledge of effective HCW protection. It monitors anti-SARS-CoV-2 immunoglobulin serum levels and collects information on symptoms of respiratory infection, work and home environment, and self-perceived SARS-CoV-2 infection risk through a standardised questionnaire survey (German Clinical Trial Registry, DRKS00021152; study protocol: https://doi.org/10.1101/2020.12.02.20242479). Starting in March 2020 as a convenience sample among employees at Hannover Medical School from the healthcare sector with direct patient contact, it has continuously been expanded. As of December 2021, the sample included a total of more than 1,000 participants (HCWs younger than 18 years were not included in CoCo). Initially, the HCWs were examined every six months and a sub-sample of 200 HCWs weekly; subsequently, frequency was adapted to general infection rates. Participation was entirely voluntary. The data base and questionnaire are administered at Hannover Medical School in accordance with German data protection law.

Sources: (48,53)

## Results

### The German health system: pandemic protection of hospital HCWs in context

The governance of HCW protection must be placed in the context of Germany’s social health insurance system, based on partnership governance of statutory health insurance funds and physicians’ associations and nearly universal health coverage (46,47). It is also shaped by EU law and health workforce regulation (6). The recent Country Health Profile (47) highlighted that Germany was relatively well prepared in terms of infrastructures and resources. It spends a greater proportion of its GDP on health (11.7%) than any other EU country (47: p.3) and health workforce staffing levels are among the highest in OECD countries (3: Figure 3.1). During the first wave of the pandemic health workforce capacity was scaled-up through a number of measures (39,47), yet HCW shortage still remained a major problem. Köppen et al. concluded from their review, that health workforce planning was limited in the pandemic response plan and ‘actions during the peak of the pandemic varied considerably across states’ (54). HCW protection policy focused on physical protection, vaccination prioritisation of HCWs and bonus payment for frontline HCWs (12).

The German hospital sector has high resources and political power. The pre-COVID-19 intensive care unit (ICU) capacity was already the highest among EU countries and quickly expanded by 20% after the start of the pandemic (47: Figure 19). For the HCWs, this expansion reinforced the problems of the pre-pandemic conditions of generally ‘high workloads in ICU and other wards, as well as nursing shortages, reflected in low rates of nurses per ICU bed’ (47,55). Similarly, minimum requirements of nurse staffing levels in intensive care and geriatric care were suspended between March and August 2020 and for high-maintenance areas until end of 2020 to help hospitals (47) These policy changes may have increased pressures on HCWs to ensure quality of care with less well educated staff. Policy efforts focussed on structural resources and technical equipment, including ensuring appropriate personal protective equipment (PPE) and surveillance measures especially for the hospital sector (12,39). The governance attempts towards strengthening HCW protection were nested in a system of high-quality hospital hygiene, infection control and regulatory frameworks of occupational health in the hospital sector (56).

### The organisation: operational governance and HCW protection at Hannover Medical School

The organisation generally plays an important role in the governance of HCW protection. In Germany, it has legal responsibility for infection protection of its employees (§23 *Infektionsschutzgesetz*) (57). To a large degree, the organisation is also accountable for surveillance and for health workforce planning and deployment. Weak governance efforts on the system level reinforce the responsibility of the organisation. This situation was observed in Germany during the pandemic (54). Shifting responsibility for operational governance to the organisational level creates flexibility and high variation between organisations, but also opens new opportunities for investing in HCW protection and preparedness.

Hannover Medical School is placed in the category of the largest German hospitals that are legally obliged to provide emergency treatment and may therefore be better equipped and prepared for maintaining flexibility and resilience during the pandemic. As an academic medical centre it can also draw on first-hand research and knowledge production. In 2020, when the CoCo Study was initiated, routine capacities of hospital beds accounted for 1,520 beds and 7,500 employees, including 3,100 HCWs (head counts) (53). COVID-19 protection management rested on two major strategies, the strengthening of surveillance management and provision of personal protective equipment (PPE) and the establishment of a new governance board, the Corona Task Force.

> ‘During the national ‘lockdown’ [spring 2020], PPE and infection control protocols at Hannover Medical School were ramped up hospital-wide. An interdisciplinary ‘corona task force’ was set up to coordinate clinical approaches regarding COVID-19 and to inform hospital staff via regular emails on the number of treated COVID-19 patients in our hospital and all measures and policies related to the pandemic. Comparably few COVID-19 cases occurred in our region and were treated in our hospital… Starting from the end of March 2020, hospital staff can undergo on-site SARS-CoV-2 nasopharyngeal swab testing when an infection is suspected’ (53).

Surveillance was organised in collaboration with the occupational health service and the hospital hygiene unit and supported by volunteers, especially medical students. The Corona Task Force was established as an ad-hoc governing body. It focuses on medicine and is legally bound to the pandemic guidelines of Lower Saxony and integrated into the Hospital Emergency Operational Management (*Krankenhauseinsatzleitung*), a wider coordination and leadership body. Appointment criteria and composition were not fully transparent; gender equality law and organisational guidelines were bypassed. As of February 2022, the Task Force was led by two male physicians; notably, the Equal Opportunity Officer of Hannover Medical School was not part of the Task Force.

Key issues of the Task Force were capacity building for COVID-19 patient care and allocation of staff according to the pandemic situation and ad-hoc needs of routine and emergency care provision. During the lockdown periods, childcare facilities were kept open, aiming to support essential staff. Similarly, psychological support services for HCWs were maintained during the pandemic, but the provision was not adequately scaled-up and adapted to new emergent needs of the HCWs.

A third pillar of HWR protection emerged in February 2021, when HCWs were included in a national vaccination prioritisation plan and vaccines were available for Lower Saxony. Hannover Medical School established their own vaccination centre, supported by the occupational health service unit and many volunteers. Vaccination was offered during work hours to all employees and students regardless of their involvement in patient care.

### Medically approved COVID-19 risk

Medical indicators prove effective organisational management of HCW protection. Analyses of serum tests revealed an infection risk of the HCWs that largely remained in the range of that of the general population of the region, although many HCWs were involved in emergency services and care for COVID-19 patients. More specifically, during the first peak of the COVID-19 pandemic (March–April 2020) anti-SARS-CoV-2 IgG prevalence was less than 1% at baseline among the participants. However, participants estimated their ‘personal likelihood of having had a SARS-CoV-2 infection with a mean of 21% [median 15%, interquartile range (IQR) 5–30%]’ (49, see also 48). Continual follow-up tests during the second wave confirmed an overall low to very low prevalence of COVID-19 infections among the HCWs (1.4% PCR positive; 0.6% silent seroconversion) (unpublished data, CoCo Study).

Most recently, during the Omicron COVID-19 wave, the Task Force reported that Hannover Medical School ranked among the top three university hospitals with the lowest ratio (0.1% of all employees) of COVID-19 quarantined employees (internal newsletter communication, 25 January 2022). These data suggest that Hannover Medical School was able to keep a very low infection rate among their HCWs throughout the entire period of the pandemic.

### Individual perceptions of risk, fear and protection: findings from the survey

Our sample comprised approximately one third of the total HCW workforce stock at Hannover Medical School, covering a wide range of health professions and occupational groups. Nurses (40%) and physicians (28%) accounted for the vast majority. About 46% of the HCWs had direct contact with COVID-19 patients. About three quarters of the respondents were women, mirroring the overrepresentation of women in the health workforce, especially in nursing [58]. Table 1 below provides further details.

**Table 1.** Composition of the sample of HCWs at Hannover Medical School

| *Items, selected* | *Sample, second wave* | *Valid % of respondents* |
| --- | --- | --- |
| *Gender* | female<br>male | 73.2<br>26.7 |
| *Workplace* | emergency room<br>ward<br>ICU<br>theatre room<br>ambulance<br>others | 10.8<br>24.6<br>17.5<br>21.3<br>11.2<br>14.6 |
| *Profession* | physicians<br>nurses<br>medical assistants<br>others | 28.4<br>39.6<br>8.7<br>23.3 |
| *COVID-19 care confirmed* | No<br>yes<br>no information | 50.4<br>45.0<br>4.6 |
| *COVID-19 care with/ without protection* | no contact<br>contact with full protection<br>contact not/not fully protected | 53.4<br>25.9<br>20.0 |

Source: CoCo Study, own calculations

More than half of the respondents (58%) estimated their own infection risk – operationalised as likelihood of having been infected – higher than 5% and 12.7% even higher than 30% (Table 2) [median=10%(IQR 1–20), mean=15.2±19.4%].

**Table 2.** Estimated likelihood of COVID-19 infection

| *Percentage of COVID-19 risk, grouped* | *Estimation of risk\*, valid percentage of respondents* |
| --- | --- |
| 0 | 23.5 |
| 0.1–5 | 18.6 |
| □ 5–10 | 24.8 |
| □ 10–20 | 10.9 |
| □ 20–30 | 9.3 |
| □ 30–100 | 12.7 |

\* Item 26 (translated). How high do you estimate the likelihood that you have already been infected?
Source: CoCo Study, own calculations

The vast majority (68.4%) expressed ‘some’ to ‘very strong’ fear of infection at the workplace’. The infection risk in the private sphere was estimated lower (56.7%), but remained an important matter of concern (Table 3).

**Table 3.** Perceptions of fear of infection at the workplace and in the private sphere, valid percentage of participants

| *5-point*<br>*likert-scale* | *Fear of infection,*<br>*Workplace\** | *Fear of infection,*<br>*private+* |
| --- | --- | --- |
| none | 4.0 | 5.8 |
| little | 27.6 | 37.3 |
| some | 42.4 | 45.8 |
| strong | 19.7 | 9.8 |
| very strong | 6.3 | 1.1 |

\* Item 33 (translated). Are you afraid of getting infected with SARS-CoV-2 in your workplace environment?
+ Item 34 (translated). Are you afraid of getting infected with SARS-CoV-2 in your private environment?
Source: CoCo Study, own calculations

Table 4 provides a comparative overview of the estimations of individual protection behaviour in the private sphere (operationalised as ‘social distancing’), the perceptions of protective measures by the employer and the compliance with protective workplace measures. More than half (60%) of the respondents estimated their own protective behaviour in the private sphere as strong to very strong and on average higher than required by law; only 1% showed ‘little’ and nobody ‘no’ compliance with protective measures. However, only less than half (49%) of the participants felt strongly protected by the employer and 12% even perceived little to no protective workplace measures. The compliance with protective measures at the workplace received overall positive rankings, yet 5% expressed negative perceptions.

**Table 4.** Individual protection behaviour and workplace measures, perceptions, valid percentage of participants

| *5-point*<br>*likert-scale* | *Estimation of own*<br>*protective behaviour,*<br>*private sphere\** | *Perception of*<br>*protective measures*<br>*of employer+* | *Perception of compliance*<br>*with protective*<br>*measures, workplace#* |
| --- | --- | --- | --- |
| none | 0.0 | 1.5 | 0.5 |
| Little | 1.0 | 10.2 | 4.2 |
| some | 39.0 | 39.3 | 24.4 |
| strong | 40.7 | 44.2 | 60.5 |
| very strong | 19.0 | 4.6 | 10.0 |

\* Item 37 (translated). Do you comply with social-distancing guidelines in your leisure time?
+ Item 41 (translated). Do you feel protected by the infection protection measures taken by your employer?

Further analyses and identification of group-specific patterns of perceptions were limited to single regression, because the strong intersections of the selected variables excluded multiple regression operations. The findings revealed that gender has no significant effect on any of the items selected for our analysis. Female and male HCWs estimated their personal infection risk and workplace protective measures similar and expressed similar perceptions of fear. However, nurses and ‘others’ estimated their own risk of having been infected higher than physicians (p=0.004). Nurses also showed the highest level of fear of infection in the private sphere compared to the other two occupational categories (p=0.004).

Contact with COVID-19 patients, no matter whether protected or not (fully) protected, had no significant effect on the estimations of risks and the compliance with protective behaviour at the workplace. However, HCWs with COVID-19 patient contact expressed higher levels of fear of infection both at the workplace and in the private sphere (p<0.001). No significant correlation could be found between full protection during COVID-19 patient care and the estimations of a personal infection risk, but contact with COVID-19 patients without protection correlated with lower estimations of protective organisational measures (p=0.001).

## Discussion

Our analysis reveals a strong mismatch between personal risk estimations and fear of infection on the one hand, and a very low medically approved infection rate coupled with strong prevention measures at Hannover Medical School on the other hand. A mean personal risk estimate of 15% against only 1% of medically approved infections, largely in the range of the general population, illustrates the different realities of individual ‘subjective’ perceptions and ‘objective’ technical measurements and organisational conditions. An overestimation of having acquired COVID-19 was already reported during the first wave, where participants estimated their personal risk with a mean of 21%, while medical tests revealed an infection rate of 1% (49). Comparing first and second wave data shows a decrease in personal risk estimations. However, the mismatch between individual and technical ‘realities’ remains strong, despite a stable trend of very low medically approved infections among HCWs over time.

The ‘dual pandemic’ approach, introduced by the European Office of the WHO (9), may help us to disentangle the different realities and to understand the complexity of physical and mental health threats of COVID-19 and its relevance for health workforce governance. The medical-technical measurements provide the best available evidence of physical health risks, which may help to estimate a temporary drop-out of infected (sick and quarantined) HCWs during the pandemic and the related surge capacities. The individual perceptions are the most relevant indicators in relation to stress of HCWs and new threats to their mental health and wellbeing. These indicators may help manage (mental) health protection and social support service more effectively and estimate middle-to long-term effects in health labour market development and workforce planning. They should therefore be included in future health workforce and pandemic preparedness plans on macro- and meso-levels of governance (4).

Fear of infection at the workplace may spill over to other areas of life and create a spiral of stress. Participants expressed higher fear of infection compared to the actual risks in relation to both the workplace and the private sphere. Contact with COVID-19 patients at the workplace seemed to increase fear of infection, regardless of being protected or not protected during patient care. This pattern might also impact negatively in the perception of protective workplace measures. Feeling poorly protected and cared for by the organisation are important sources of stress, which may be correlated to burn-out of HCWs, as a recent study with UK nurses and midwives during the first wave of the pandemic shows (59). Individual perceptions and concerns need greater attention in health workforce research and policy, no matter whether there is a technical ‘objective’ reason or not.

Notably, women account for the vast majority of HCWs and many of them are nurses, who showed the highest levels of fear of infection and risk estimations in our study. Gender differences, reported in some studies in relation to higher levels of infection (60) and/or fear among female HCWs (48,49), might therefore be an interconnected effect of a female-dominated nursing profession as the largest health labour market segment. The results call for greater attention to the perceptions and needs of both nurses and female HCWs during the pandemic. The literature highlights fear and exhaustion of nurses and some studies report higher infection rates (30,31,61,62). There is also growing evidence of an increase in gender inequalities in the health workforce and in leadership positions during the pandemic (22,35,63,64,65), confirming previous observations that new emergent management structures may bypass gender equality guidelines on the organisational level (66). It should be noted that other factors not measured by our data, e.g. the migration status, might intersect with the professional status and sex/gender categories (23,24,67). Future pandemic preparedness plans and COVID-19 management should improve both inclusion of nurses in decision-making bodies and female leadership.

Finally, our optimal-case scenario shows low levels of infection compared to other countries [3,16,18,24,26,68], as well as within Germany [17,47,57,69]. These optimal conditions may not easily be translated to other contexts, yet there are some important lessons emerging from our case study in relation to health workforce governance and new approaches to organisational protection measures. Most importantly, the findings reveal that protecting HCWs from COVID-19 infection is not enough. HCWs overestimate their personal risk even under conditions of rather optimal protection at the workplace, thus creating stress and new threats to the individual health of HCWs and to health labour market development (25). These threats may be preventable, at least to some degree, through innovation in the governance of pandemic preparedness and health workforce protection (10,36,52), that take the ‘dual pandemic’ (9) dimension of COVID-19 and the ‘human face’ of the health workforce (44) more systematically into account.

### Limitations

Our study reveals important gaps in HCW protection and health workforce governance. However, it has several limitations, which have been described in relation to the clinical part of the study (48,49). To summarise the major arguments: data on anti-SARS-CoV-2 IgG is only partially representative for Hannover Medical School and we do not know the source of infection in anti-SARS-CoV-2 IgG-positive HCWs (49). More specifically related to our selected survey data, the respondents might be biased; employees who are more concerned about their health and a COVID-19 infection might have been more interested in the study than those who do not care about potential health risks. We also do not know how the experience of an organisational environment characterised by low infection rates intersects with the individual sphere, and how different sources of information impact in individual perceptions. Further research and qualitative methodology would be necessary to provide in-depth information. Finally, the lessons that can be learned from an ‘optimal-case scenario’ in relation to individual perceptions and institutional/organisational conditions of HCW protection are generally limited and must be viewed with caution, because cross-country comparative data and in-depth organisational comparison are lacking. Our results may help to highlight the need for, and benefit of more comprehensive research and policy investigation into mental health of HCWs.

## Conclusions

We set out to make ‘blind spots’ of health workforce protection during the pandemic visible and to highlight the need for greater attention to the individual perceptions and mental health requirements of HCWs. Our findings reveal a strong mismatch between technical measures and individual perceptions of HCWs. Applying a ‘dual pandemic’ approach to COVID-19 (9) opens new opportunities to explore this mismatch in more detail and develop governance approaches, that respond more effectively to health workforce needs and resilience (2,8,12,39). Notwithstanding the importance of country-specific contexts and the privilege of a high-resourced healthcare system and low-risk setting, we believe that our case study may support much needed investment in the health workforce and help build back better and fairer after the pandemic. Our findings highlight that improving health workforce funding and planning are not enough, but greater attention to mental health and wellbeing of HCWs could make a difference.

What lessons for pandemic recovery plans and investment in the health workforce?

- Invest in mental health and HCW’s wellbeing, improve research evidence and create new mental health and social support services as part of future pandemic plans.
- Improve transparency and develop information and risk communication as part of COVID-19 infection prevention and HCW protection management.
- Establish inclusive multi-professional governance models based on participatory governance and strengthen the role of nurses in HCW protection and pandemic management.
- Strengthen women’s inclusion and female leadership in future pandemic plans to improve mental health protection and ensure equal opportunities in all areas of decision-making.

## Data Availability

All data produced in the present study are available upon reasonable request to the authors

## Declarations

### Data availability

CoCo datasets can be obtained from at reasonable request.

## Ethics statement

Ethical approval for the survey was obtained from Hannover Medical School; https://doi.org/10.1101/2020.12.02.20242479; 8973_BO_K_2020.

## Author contributions

EK and AD-J had the idea for the paper, developed the conceptual approach, wrote the first draft and revised the manuscript; GB, AD-J and CH conceptualised the CoCo Study; AC collected the data; SH carried out statistical analyses; all authors have read and approved the final version of the manuscript.

## Acknowledgements

We thank the participants in the survey for sharing their experiences and Marianne Richter for supporting the statistical analysis.

## References

1 WHO. Year of the Health and Care Workers 2021: Protect. Invest. Together. Geneva: WHO (2021); https://www.who.int/campaigns/annual-theme/year-of-health-and-care-workers-2021 (accessed 5 February 2022).

2 Zapata T, Buchan J, Azzopardi-Muscat N. The health workforce: central to an effective response to the COVID-19 pandemic in Europe. Int J Health Plann Manage. (2021) 36(S1):9–13.

3 European Commission (EC). State of Health in the EU. Companion Report 2021. Luxemburg: European Union (2021); https://ec.europa.eu/health/sites/default/files/state/docs/2021_companion_en.pdf (accessed 8 January 2022).

4 European Commission (EC). Supporting mental health of the health workforce and other essential workers. Fact sheet accompanying the Opinion by the Expert Panel on Effective Ways of Investing in Health (EXPH). Luxemburg: European Union (2021); https://op.europa.eu/en/publication-detail/-/publication/5c342879-2fb9-11ec-bd8e-01aa75ed71a1 (accessed 8 January 2022).

5 EUPHA-EHMA. Strengthening the EU’s competitiveness requires an increased investment in the workforce and governance. Advocacy Statement (2022); https://eupha.org/advocacy-by-eupha (accessed 3 February 2022).

6 Panteli D, Maier CB. Regulating the health workforce in Europe: implications of the COVID-19 pandemic. Hum Resour Health. (2021) 19:80.

7 Williams GA, Scarpetti G, Bezzina A, Vincenti K, Grech K, Kowalska-Bobko I, et al. How are countries supporting their health workers during Covid-19? Ensuring sufficient workforce capacity. Eurohealth. (2020) 26(2):58–62.

8 Azzopardi-Muscat N. A public health approach to health workforce policy development in Europe. Eur J Public Health. (2020) 30(Suppl. 4):iv3–iv4.

9 Azzopardi-Muscat N. Interview. WHO: COVID-19 will be a ‚dual pandemic’ – physical and mental. Brussels: EU-Observer (2021); https://euobserver.com/coronavirus/153088 (accessed 10 January 2022).

10 Buchan J, Williams G, Zapata T. Governing health workforce responses during COVID-19. Eurohealth. (2021) 27(1):41–48.

11 Kluge H. Statement COVID-19: taking stock and moving forward together. Copenhagen: WHO Regional Office for Europe, 2020; https://www.euro.who.int/en/media-centre/sections/statements/2020/statement-covid-19-taking-stock-and-moving-forward-together; (accessed 5 February 2022).

12 Kuhlmann E, Brinzac MG, Burau V, Correia T, Ungureanu M-I. Health workforce preparedness and protection during the COVID-19 pandemic: a tool for the rapid assessment of European Union countries. Eur J Public Health. (2021) 31(Suppl. 4):iv14– iv20.

13 WHO. Supporting the mental health needs of the health workforce. Copenhagen: WHO (2021); https://apps.who.int/iris/bitstream/handle/10665/340220/WHO-EURO-2021-2150-41905-57496-eng.pdf (accessed 20 January 2022).

14 Baptista S, Teixeira A, Castro L, Cunha M, Serrão C, Rodrigues A, Duarte I. Physician burnout in primary care during the COVID-19 pandemic: a cross-sectional study in Portugal. J Prim Care Community Health. (2021)12:1–9.

15 De Wit K, Mercuri M, Wallner C, Clayton N, Archambault P, Richie K, et al. Canadian emergency physician psychological distress and burnout during the first 10 weeks of COVID-19: a mixed-methods study. JACEP Open. (2020)1:1030–1038.

16 Ferland L, Carvalho C, Comes Dias JG, Lamb F, Adlhoch C, Suetens C, et al. Risk of hospitalization and risk of death for healthcare workers with COVID-19 in nine European Union/European Economic Area countries, January 2020–January 2021. J Hosp Infect. (2022) 119:170–174.

17 Fischer-Fels J. Gesundheitspersonal und COVID-19. Infektionszahlen nehmen zu. Dtsch Arztebl. (2020)117(31/32):1484.

18 Guillén E, Buissonnière M, Lee CT. From lionising to protecting healthcare workers during and after COVID-19: systems solutions for human tragedies. Int J Health Plann Manage. (2021) 36(S1):20–25.

19 Hummel S, Oetjen N, Du J, Posenato E, Resende de Almeida RM, Losada R, et al. Mental health among medical professionals during the COVID-19 pandemic in eight European countries: cross-sectional survey study. J Med Internet Res. (2021) 23(1):e24983.

20 Kuhlmann E, Bruns L, Hoeper K, Richter, M., Witte T, Ernst D, Jablonka A. Work situation of rheumatologists and residents in times of COVID-19: findings from a survey in Germany. Z Rheumatol. (2021); ahead of print, DOI:10.1007/s00393-021-01081-5.

21 Lorello GR, Gautam M, Barned C, Peer M. Impact of the intersection of anaesthesia and gender on burnout and mental health, illustrated by the COVID-19 pandemic. Anaesthesia. (2021) 76(Suppl. 4):24–31.

22 Lotta G, Fernandez Pimenta D, Wenham C. Gender, race and health workers in the COVID-19 pandemic. Lancet. (2021) 397:1264.

23 Lotta G, Fernandez M, Correa M. The vulnerabilities of the Brazilian health workforce during health emergencies: analysing personal feelings, access to resources and work dynamics during the COVID-19 pandemic. Int J Health Plann Manage. (2021) 36(S1):42–57.

24 Lotta G, Nunes J, Fernandez M, Corrêa MC. The impact of the COVID-19 pandemic in the frontline health workforce: perceptions of vulnerability of Brazil’s community health workers. Health Policy Open. (2022) 3:100065.

25 Marburger Bund. Umfrage: Klinikärzte sind regelmäßig erschöpft – jeder fünfte plant Tätigkeitswechsel. News blog. (13 February 2022); https://www.marburger-bund.de/bundesverband/pressemitteilung/umfrage-klinikaerzte-sind-regelmaessig-erschoepft-jeder-fuenfte (accessed 14 February 2022).

26 Nguyen LH, Drew DA, Graham MS, Joshi AD, Guo C-G, Ma MSW, et al. Risk of COVID-19 among front-line health-care workers and the general community: a prospective cohort study. Lancet Public Health. (2020) 5(9):e475–e483.

27 Wahlster S, Sharma M, Lewis AK, Patel PV, Hartog CS, Jannotta G, et al. The coronavirus disease 2019 pandemic’s effect on critical care resources and health-care providers. A global survey. Cest. (2021) 159(2):619–633.

28 White EM, Wetle TF, Reddy A, Baier RB. Front-line nursing home staff experiences during the COVID-19 pandemic. JAMDA. (2021) 22:199–203.

29 Zhou AY, Hann M, Panagioti M, Patel M, Agius R, van Tongeren M, et al. Cross-sectional study exploring the association between stressors and burnout in junior doctors during the COVID-19 pandemic in the United Kingdom. J Occup Health. (2022) 64:e12311.

30 Buchan J, Catton H, Sfaffer F. Sustain and retain in 2022 and beyond: the global nursing workforce and the COVID-19 pandemic. Philadelphia: International Center on Nurse Migration (2022); https://www.icn.ch/system/files/2022-01/Sustain%20and%20Retain%20in%202022%20and%20Beyond-%20The%20global%20nursing%20workforce%20and%20the%20COVID-19%20pandemic.pdf (accessed 4 February 2022).

31 WHO. Frontline stories: mental health of health care workers in the COVID-19 pandemic. Geneva: WHO (2021); https://www.euro.who.int (accessed 1 February 2022).

32 Lancet. Healthcare workers owed a better future. Lancet. (2021) 397:347.

33 Campbell J, Dussault G, Buchan J, dal Poz M, Guerra-Arias M, Leone C, et al. A universal truth: no health without a workforce. Third Global Forum on Human Resources for Health, Recife, Brazil (2013); https://www.who.int/workforcealliance/knowledge/resources/GHWA-a_universal_truth_report.pdf?ua=1 (accessed 5 February 2022).

34 Shaw A, Flott K, Fontana G, Durkin M, Darzi A. No patient safety without health worker safety. Lancet. (2020) 396:1441–1442.

35 Bourgeault IL, Maier CB, Dieleman M, Ball J, MacKenzie A, Nancarrow S, et al. The COVID-19 pandemic presents an opportunity to develop more sustainable health workforces. Hum Resour Health. (2020) 18:83.

36 Greer SL, King EJ, Massard da Fonseca E. In Introduction. Edited by Greer SL, King EJ, Massard da Fonseca E, Peralta-Santos A. Coronavirus Politics. The Comparative Politics and Policy of COVID-19. Ann Arbor, MI: University of Michigan Press (2021: p.3– 33); https://doi.org/10.3998/mpub.11927713 (accessed 5 February 2022).

37 Forman R, Azzopardi-Muscat N, Kirkby V, Lessof S, Limaro Nathan N, Pastorino G, et al. Drawing light from the pandemic: Rethinking strategies for health policy and beyond. Health Policy. (2022) 126:1–6.

38 European Union Expert Panel (EU). The organisation of resilient health and social care following the COVID-19 pandemic. Opinion of the Expert Panel on effective ways of investing in Health (EXPH). Luxembourg: Publications Office of the European Union (2020).

39 Burau V, Falkenbach M, Neri S, Peckham S, Wallenburg I, Kuhlmann E. Health system resilience and health workforce capacities: comparing health system responses during the COVID-19 pandemic in six European countries. Int J Health Plann Manage. (2022) ahead of print, DOI: https://doi.org/10.1002/hpm.3160.

40 McKee M. Drawing light from the pandemic. A review of the evidence for the Pan-European Commission for Health and Sustainable Development. Copenhagen: WHO (2021); https://www.euro.who.int/data/assets/pdf_file/0015/511701/Pan-European-Commission-health-sustainable-development-eng.pdf (accessed 8 January 2022).

41 McKee M. Building back better: why we need to fix the health worker divide in Europe. Eur J Public Health. (2021) 31(4):669.

42 Rieckert A, Schuit E, Bleijenberg A, ten Cate D, de Lange W, de Man-van Ginkel Jm, et al. How can we build and maintain the resilience of our health care professionals during COVID-19? Recommendations based on a scoping review. BMJ Open. (2021) 11:e043718.

43 Sagan, A, Webb E, Azzopardi-Muscat N, de la Mata I, McKee M, Figueras J. Health systems resilience during COVID-19: lessons learned to build back better. Health Policy Series 56. Copenhagen: WHO (2021).

44 Kuhlmann E, Dussault G, Wismar M. Health labour markets and the ‘human face’ of the health workforce: resilience beyond COVID-19. Eur J Public Health. 2020;30(Suppl. 4):iv1–iv2.

45 Czypionka T, Reiss M. Three approaches to handling the COVID crisis in federal countries: Germany, Austria, and Switzerland. In Coronavirus Politics. The Comparative Politics and Policy of COVID-19. edited by Greer SL, King EJ, Massard da Fonseca e, Peralta-Santos A. Ann Arbor, MI: University of Michigan Press (2021, p.295–319); https://doi.org/10.3998/mpub.11927713 (accessed 5 February 2022).

46 Busse R, Blumel M, Knieps F, Bärnighausen T. Germany and health 1. Statutory health insurance in Germany: a health system shaped by 135 years of solidarity, self-governance, and competition. Lancet. (2017) 390:882–897.

47 OECD/European Observatory on Health Systems and Policies (OBS). Germany: Country Health Profile 2021, State of Health in the EU. Paris: OECD Publishing/ Brussels:European Observatory on Health Systems and Policies (2021); file:///C:/Users/asus/AppData/Local/Temp/Germany-CountryHealthProfile2021.pdf (accessed 14 December 2021).

48 Behrens GMN, Cossmann A, Stancov MV, Schulte B, Streeck H, Förster R, et al. Strategic anti-SARS-CoV-2 serology testing in a low prevalence setting: the COVID-19 Contact (CoCo) Study in healthcare professionals. Infect Dis Ther. (2020) 9(4): 837–849.

49 Behrens GMN, Cossmann A, Stankov MV, Witte T, Ernst D, Happle C, Jablonka A. Perceived versus proven SARS-CoC-2 specific responses in health-care professionals. Infection. (2020) 48(4):631–634.

50 Hannover Medical School, MHH Medizinische Hochschule Hannover (2022); www.mhh.de (accessed 22 February 2022)

51 Dieleman M, Shaw DM, Zwanikken P. Improving the implementation of health workforce policies through governance: a review of case studies. Hum Resour Health. (2011) 9:10.

52 Lim MYH, Lin V. Governance in health workforce: how do we improve the concept? A network-based, stakeholder-driven approach. Hum Resour Health. (2021) 19:1.

53 Jablonka A, Happle C, Cossmann A, Stankov MV, Ernst D, Scharff AZ, Behrens GMN. Protocol for longitudinal assessment of SARS-CoV-2-specific immune responses in healthcare professionals in Hannover, Germany: the prospective, longitudinal, observational COVID-19 Contact (CoCo) study. Preprint (2020); https://doi.org/10.1101/2020.12.02.20242479 (accessed 10 February 2022).

54 Köppen J, Hartl K, Maier CB. Health workforce response to Covid-19: What pandemic preparedness planning and action at the federal and state levels in Germany? Int J Health Plann Manage. (2021) 6(S1):71–91.

55 Zander B, Köppen J, Busse R. Personalsituation in deutschen Krankenhäusern in internationaler Perspektive. In Krankenhausreport 2017: Schwerpunkt Zukunft gestalten. edited by Klauber J, Geraedts M, Friedrich J, Wasem J. Stuttgart: Schattauer (2017, 61–78).

56 Robert-Koch-Institute (RKI). Commission for Hospital Hygiene and Infection Prevention (KRINKO). Berlin: RKI (2021); https://www.rki.de/EN/Content/Institute/Committees/KRINKO/KRINKO_node_en.html(accessed 18 January 2021).

57 Ärzteblatt. News. SARS-CoV-2: 33.000 Neuinfektionen bei Gesundheitspersonal im Januar. Aerzteblatt.de (2 February 2021); https://www.aerzteblatt.de/nachrichten/120743/SARS-CoV-2-33-000-Neuinfektionen-bei-Gesundheitspersonal-im-Januar (accessed 5 February 2022).

58 OECD. Health statistics. Paris: OECD (2022); https://stats.oecd.org/Index.aspx?DatasetCode=HEALTH_STAT# (accessed 18 January 2022).

59 Couper K, Murrels T, Sanders J, Anderson JE, Blake H, Kelly D, et al. The impact of COVID-19 on the wellbeing of the UK nursing and midwifery workforce during the first pandemic wave: A longitudinal survey study. Int J Nurs Stud. (2021) 127(12):104155.

60 WHO. Statement – International Women’s Day: the need to build back better, with women in the lead. Copenhagen: WHO (2021); https://www.euro.who.int/en/media-centre/sections/statements/2021/statement-international-womens-day-the-need-to-build-back-better,-with-women-in-the-lead (accessed 5 March 2021).

61 European Specialised Nurses Organisation (ESNO). Caring4Nurses Campaign. Don’t let them go. Brussels: ESNO (2022); https://www.esno.org/Caring4Nurses.html (accessed 7 February 2022).

62 ICN. International Council of Nurses COVID-19 update. Geneva: ICN (2021); https://www.icn.ch/sites/default/files/inline-files/ICN%20COVID19%20update%20report%20FINAL.pdf (accessed 16 March 2021).

63 Betron M, Bourgeault I, Manzoor M, Paulino E, Steege R, Thompson K, Wuliji T, on behalf of the Global Health Workforce Network’s Gender Equity Hub. Time for gender-transformative change in the health workforce. Lancet. (2019) 393:e25.

64 Gabster RP, van Daalen K, Dhatt R, Barry M. Challenges for the female academic during the COVID-19 pandemic. Lancet. (2020) 395:1968–1969.

65 Gupta N. Research to support evidence-informed decisions on optimizing gender equity in health workforce policy and planning. Hum Resour Health. (2019) 17:46.

66 Kuhlmann E, Ovseiko P, Kurmeyer C, Gutiérrez-Lobos K, Steinböck S, von Knorring M, et al. Closing the gender leadership gap: a multi-centre cross-country comparison of women in management and leadership in academic health centres in the European Union. Hum Resour Health. (2017) 15:2.

67 WHO. Putting equity at the heart of COVID-19 recovery. Copenhagen: WHO (2021); https://www.euro.who.int/en/media-centre/events/events/2021/04/world-health-day-2021/news/news/2020/04/world-health-day-putting-equity-at-the-heart-of-covid-19-recovery (accessed 5 February 2022).

68 Gazaro G, Clari M, Ciocan C, Grillo E, Mansour I, Godono A, et al. COVID-19 infection and diffusion in a large university-hospital in northwest Italy. Med Lav. (2020) 111(3):184–194.

69 Pfenninger EG, Christ P, Neumüller M, Dinse Lambracht A. Beurteilung des Infektionsrisikos durch SARS-CoV-2 für medizinisches Personal – Erkenntnisse aus der Praxis. Bundesgesundheitsblatt. (2021) 64:304–313.

